# Rationale and Design of RECOVER-ENERGIZE: A Platform Clinical Trial of Interventions for Exercise Intolerance With and Without Post-exertional Malaise in Long COVID

**DOI:** 10.64898/2026.06.02.26354455

**Authors:** Janna Friedly, Lucinda Bateman, Lisa G. Berdan, Richard Casaburi, Nathaniel Erdmann, G. Michael Felker, Nilda Itchon-Ramos, Steven J. Keteyian, Neil MacIntyre, Lisa O’Brien, Craig Reist, Harry B. Rossiter, Adam Silverstein, Emily Taylor, Helena Pike Welch, N David Yanez, Kanecia O. Zimmerman, Barry Make

**Affiliations:** Department of Rehabilitation Medicine, University of Washington, Seattle, WA; Bateman Horne Center, Salt Lake City, UT; National Heart, Lung, and Blood Institute, National Institutes of Health, Bethesda, MD; Respiratory Research Center, The Lundquist Institute for Biomedical Innovation at Harbor-UCLA Medical Center, and Division of Respiratory and Critical Care Physiology and Medicine, Harbor-UCLA Medical Center, Torrance, CA; Department of Medicine, University of Alabama at Birmingham, Birmingham, AL; Duke Clinical Research Institute, Duke University School of Medicine, Durham, NC; Henry Ford Health, Division of Cardiovascular Medicine, Detroit, MI; Department of Medicine, Duke University School of Medicine, Durham, NC; RECOVER Patient, Caregiver, or Community Representative, Durham, NC; Social, Statistical, and Environmental Sciences, RTI International, Research Triangle Park, NC; Department of Biostatistics & Bioinformatics, Duke University School of Medicine, Durham, NC; Department of Pediatrics, Duke University School of Medicine, Durham, NC; Department of Medicine, National Jewish Health, Denver, CO

**Keywords:** Post-exertional malaise, Cardiopulmonary rehabilitation, Exercise intolerance, Long COVID, Post-acute sequelae of SARS-CoV-2 infection

## Abstract

**Introduction:** A prominent symptom of post-acute sequelae of SARS-CoV-2 infection (i.e., Long COVID) is exercise intolerance with or without post-exertional malaise (PEM). PEM is characterized by the worsening of both symptoms and function following even minor physical or mental exertion, with symptoms typically worsening 12 to 48 hours after activity and lasting for days or even weeks. Individualized, supervised cardiopulmonary rehabilitation is considered a safe and effective intervention for many cardiac and pulmonary conditions, and has been effective in gradually improving function in previously hospitalized and nonhospitalized patients with severe COVID-19. While traditional cardiopulmonary rehabilitation approaches appear helpful in some situations, the exercise intolerance symptoms experienced by many individuals with Long COVID may require a different approach, especially when attempts to increase physical activity result in PEM. No clear consensus exists on the optimal treatment of PEM, and no major studies have evaluated the efficacy in individuals with Long COVID of either carefully supervised, individualized cardiopulmonary rehabilitation programs for exercise intolerance without significant PEM or activity pacing interventions designed to treat or prevent PEM.

**Methods and Analysis:** The Researching COVID to Enhance Recovery Clinical Trials (RECOVER-CT) initiative funded by the National Institutes of Health (NIH) included a prospective, multicenter, randomized controlled platform trial (RECOVER-ENERGIZE) designed to assess two interventions in patients with Long COVID and exercise intolerance: (1) cardiopulmonary rehabilitation for patients without significant PEM and (2) structured activity pacing to prevent or reduce PEM in participants who experience the symptom. The intervention duration will be 12 weeks. The primary endpoints for the trial include the Endurance Shuttle Walk Test as a measure of endurance capacity for the cardiopulmonary rehabilitation intervention and a modified version of the DePaul Symptom Questionnaire–Post-Exertional Malaise for the pacing intervention. Assessments will be completed at baseline, middle of intervention, end of intervention, and 12 weeks after completion of the intervention, and include physical performance measures and patient-reported surveys.

**Ethics and Dissemination:** The RECOVER-ENERGIZE trial protocol has been approved by an institutional review board (Advarra), and written informed consent will be obtained from all participants prior to enrollment. The trial is registered on ClinicalTrials.gov (NCT06404047). Formally assessing PEM and developing a structured activity pacing intervention delivered by local pacing coaches are novel features of this trial. Results will be disseminated through peer-reviewed publications, presentations at scientific conferences, and communication with participants, patient advocacy organizations, and the broader Long COVID community. De-identified participant data will be made available through the NIH RECOVER data repository in accordance with NIH data-sharing policies. If successful, this protocol will provide accessible tools that clinicians can use to address exercise intolerance and PEM in patients with Long COVID.

**Trial registration:** ClinicalTrials.gov – Platform: NCT06404047; Appendix A: NCT06404060; Appendix B: NCT06404073. Registered on May 6, 2024.

**Strengths and limitations of this study:**

- RECOVER-ENERGIZE is a large, multicenter, randomized controlled platform trial that stratifies participants by PEM status, separately evaluating cardiopulmonary rehabilitation in those without significant PEM and structured activity pacing in those with PEM, while mitigating the risk of exertional harm.
- The structured activity pacing intervention is novel and has not previously been tested in a randomized trial in Long COVID. Its coach-delivered, video-conference format is designed to be easily implemented and scalable across diverse clinical settings.
- Patient, caregiver, and community representatives were integrally involved throughout protocol development, shaping eligibility criteria, intervention design, and selection of outcome measures, which strengthens the relevance of the trial to the Long COVID community.
- The trial combines a performance-based measure of endurance capacity (the Endurance Shuttle Walk Test) with a modified, PEM-specific patient-reported instrument (mDSQ-PEM). However, the nature of the interventions precludes blinding of participants and providers, and several key outcomes rely on self-report, which may introduce bias.

## BACKGROUND

Post-acute sequelae of SARS-CoV-2 infection (PASC, also known as Long COVID) is a chronic condition that can occur regardless of acute disease severity, affecting up to 20% of infected individuals across all demographics.^1–3^ It affects nearly every organ system, with more than 200 individual symptoms, including new-onset fatigue, cognitive difficulties, shortness of breath, anxiety, depression, dizziness, and arrhythmias.^4^ These prolonged symptoms are associated with substantial short- and long-term individual and societal costs, including inability to work and healthcare expense.^5,6^ Effective prevention and treatment strategies for Long COVID remain an urgent public health priority.

Exercise intolerance, characterized by reduced capacity for physical activity due to dyspnea, early muscle fatigue, or the sensation of tiredness during exercise, is one of the most common symptoms of Long COVID and can have long-term effects on quality of life and survival.^7–10^ Post-exertional malaise (PEM), sometimes referred to as post-exertional neurological symptoms, is a related symptom that involves worsening of physical, cognitive, and emotional function after minimal exertion, often peaking 12–48 hours after activity and lasting days to weeks. Patients who experience PEM often have a markedly reduced ability to engage in exercise or even perform routine activities of daily living following physical, mental, cognitive, or emotional exertion.^11–12^ PEM resembles activity intolerance in myalgic encephalomyelitis/chronic fatigue syndrome (ME/CFS) and may result from nervous system, immune, and energy metabolism dysregulation. Graded exercise has been shown harmful for individuals with PEM, warranting alternative rehabilitation approaches.^13^

Cardiopulmonary rehabilitation is highly effective and scalable to improve exercise capacity and quality of life in various chronic diseases and may benefit some patients with Long COVID.^14–16^ However, traditional cardiopulmonary rehabilitation may need modification especially when attempts to exercise result in PEM.^17^ Pacing, a strategy to limit activity within an “energy envelope,” is an established strategy for PEM associated with ME/CFS and other chronic fatiguing illnesses^18–20^ and is emerging as a promising intervention to reduce the likelihood, severity, and duration of Long COVID–related PEM.

There is currently no clear consensus on the optimal treatment of PEM, and no clinical trials have systematically evaluated the efficacy of either carefully supervised, individualized physical conditioning programs that account for PEM and other symptoms, or activity pacing interventions designed to prevent PEM in Long COVID. As part of the National Institutes of Health (NIH)–funded Researching COVID to Enhance Recovery Clinical Trials (RECOVER-CT) initiative, the RECOVER-ENERGIZE platform trial will evaluate:

1. A 12-week personalized cardiopulmonary rehabilitation program for patients with exercise intolerance without significant PEM (“Appendix A”).
2. Structured pacing intervention for patients with significant PEM (“Appendix B”). Outcomes include changes in exercise tolerance and patient-reported measures, aiming to inform optimal rehabilitation strategies for Long COVID.

## METHODS

### Participants

To be included in the overall platform trial, individuals must be ≥18 years of age; have a previous suspected, probable, or confirmed SARS-CoV-2 infection as defined by the Pan American Health Organization^21^; and have a self-reported limitation to physical activity due to the presence of symptoms such as fatigue, shortness of breath, and/or PEM that has persisted for at least 12 weeks and is present at the time of consent. Participants are required to provide written informed consent, complete questionnaires and outcome assessments, and participate in assigned intervention or control and study visits, whether remote, hybrid, or in-person. The study protocol has been approved by the RECOVER CT Data Safety Monitoring Board, Advarra Institutional Review Board (Pro00068561), and each site’s local institutional review board if required by their institution. Due to the nature of the interventions, this study is not blinded to participants or providers except for the study staff who conduct the Endurance Shuttle Walk Test (ESWT) for Appendix A.

Individuals will be excluded from participating if they have been diagnosed with known active acute SARS-CoV-2 infection ≤4 weeks prior to the consent, if they have a prior diagnosis of ME/CFS not related to SARS-CoV-2 infection, or if they use (or used within the last 14 days) a formal program incorporating one or more of the ENERGIZE interventions or a similar intervention to treat the underlying condition.

#### Additional Appendix A Inclusion and Exclusion Criteria

There are no additional inclusion criteria for Appendix A (cardiopulmonary rehabilitation). Additional exclusion criteria for Appendix A include known pre-existing postural orthostatic tachycardia syndrome, not related to SARS-CoV-2 infection; known uncontrolled hypertension (blood pressure ≥ 160/100 mmHg at rest); any of the following within 4 weeks of enrollment: acute myocardial infarction, unstable angina, uncontrolled arrhythmias causing symptoms or hemodynamic compromise, acute myocarditis or pericarditis, uncontrolled acutely decompensated heart failure (acute pulmonary edema), acute pulmonary embolism, suspected dissecting aneurysm, severe hypoxemia at rest, a thromboembolic event, or any acute or chronic disorder that may affect exercise capacity or be aggravated by exercise (e.g., infection, exercise-induced syncope, thyrotoxicosis); being unable to cooperate; the presence of orthostatic hypotension as defined by a selection of ≥8 on question 1 or ≥9 on question 3 of the Orthostatic Hypotension Daily Activity Scale from the modified Orthostatic Hypotension Questionnaire (mOHQ)^22^; having engaged in purposeful moderate or greater intensity exercise with the intent to improve one’s health 2 or more times per week over the 30 days prior to informed consent; or inability to walk. With input from patient advocates and in recognition of the importance of avoiding the potential harm of an exercise intervention in patients with significant PEM, we excluded patients who reported baseline moderate to severe self-reported PEM using a modified version of the DePaul Symptom Questionnaire–Post-Exertional Malaise (mDSQ-PEM) validated in patients with PEM.^23^ The mDSQ-PEM measures the frequency, duration and severity of PEM-related symptoms including fatigue, cognitive impairment, and pain within the last 6 months, modified for this study to include different look-back periods (see details in Primary Outcome Measures section below). This questionnaire also assesses triggers of symptom exacerbation to assess whether minimal effort, exercise, or mental activity can lead to symptom exacerbation, how long it takes to recover after exertion (e.g., hours, days, or longer) and how PEM affects daily activities, physical endurance, and cognitive performance (see Supplemental Material for questionnaire).

We also excluded patients if they experienced PEM during the exercise involved in the screening process. We defined moderate or severe PEM by the following:

- a score of 2 or greater for *both* frequency *and* severity for any of the first 5 questions on the Screening mDSQ-PEM *and* answer of ‘YES’ to either item 7 or 8 on the Screening mDSQ-PEM, *and* response of > 14 hours in item 9; *or*
- a score of 3 or greater on *any severity question* (regardless of frequency) *and* answer of ‘YES’ to either item 7 or 8 on the Screening mDSQ-PEM, *and* response of > 14 hours in item 9; *or*
- any new score of 3 or greater on *any of the severity questions* on the mDSQ-PEM 48-72 hours following the Screening Incremental Shuttle Walk Test (ISWT).^24^

Individuals who are screened for Appendix A but are found to have moderate or severe PEM and are therefore not eligible for the rehabilitation intervention may be offered screening for enrollment in Appendix B (the structured pacing intervention), provided they meet Appendix B eligibility criteria.

#### Additional Appendix B Inclusion and Exclusion Criteria

To be eligible to participate in Appendix B (pacing intervention trial), individuals must identify new PEM following a SARS-CoV-2 infection that has persisted for at least 12 weeks and is still present at the time of consent and report a score of 2 or greater for both frequency *and* severity for any of the first 5 questions on the Screening mDSQ-PEM *and* answer of ‘YES’ to either item 7 or 8 on the Screening mDSQ-PEM, *and* response of >14 hours in item 9; *or* score of 3 or greater on any severity question (regardless of frequency) *and* answer of ‘YES’ to either item 7 or 8 on the Screening mDSQ-PEM, *and* response of >14 hours in item 9.

### Interventions

#### Appendix A: Cardiopulmonary Rehabilitation

Cardiopulmonary rehabilitation combines elements of standard cardiac and pulmonary programs to address the mixed causes of exercise intolerance in Long COVID. This hybrid approach is tailored to improve tolerance by incorporating strategies from both disciplines. Importantly, the ENERGIZE cardiopulmonary rehabilitation intervention is not graded exercise therapy. Unlike graded exercise therapy, which follows a fixed, progressive increase in exercise duration and intensity, the ENERGIZE rehabilitation program is individualized, symptom-limited, and closely monitored for the development of PEM. Exercise intensity and duration are adjusted based on each participant’s symptoms and tolerance, and participants are instructed to modify or stop activity if PEM symptoms occur.

On day 1, all participants undergo an in-person baseline assessment by a trained clinician to determine starting levels for lower-extremity aerobic exercise. A therapist (physical, occupational, rehabilitation, respiratory, or exercise) also evaluates strength, balance, range of motion, and flexibility to guide individualized programming, which may begin with recumbent exercise if needed. The Short Physical Performance Battery^25^ is administered by trained therapists to standardize assessment. Site rehabilitation staff receive training on PEM recognition and management to ensure programs are adapted promptly if symptoms arise. Participants who develop significant PEM during the rehabilitation intervention are discontinued from the Appendix A intervention, as continued exercise in the setting of PEM could be harmful. These participants may be offered screening for enrollment in Appendix B (the structured pacing intervention) if they meet the eligibility criteria and after completing a 90-day washout period.

The program runs for 12 weeks, with a minimum of two sessions weekly, increasing to three as tolerated. During week 1 and at least one session in week 12, participation must be in-person; remaining sessions may be in-person, remote synchronous, or asynchronous. Remote sessions are permitted only after participants demonstrate independence in prescribed exercises. Choice of delivery format is based on participant needs (e.g., transportation, work schedule) and program resources (e.g., remote monitoring capability). All participants receive home exercise instructions emphasizing PEM self-monitoring and modification or stopping of activity if symptoms occur.

In-person sessions offer group social support and may include heart rate, oxygen saturation, and blood pressure monitoring. Each session is individualized from baseline findings, symptoms, and progress, and generally includes:

1. Aerobic exercise: Progressing to ≥30 min as tolerated, with initial duration tailored to ability. Target intensity: Borg Rating of Perceived Exertion/Perceived Dyspnea score of 5–6 (scale 1–10) and heart rate > 60% predicted maximum, adjusted for symptoms. Modalities may include walking, treadmill, or upright/recumbent cycling, selected according to clinical factors and equipment availability.
2. Education: Exercise fundamentals, PEM recognition, self-management strategies, stress/anxiety/depression management, and nutrition/weight control.
3. Strength and flexibility training: Individually prescribed by rehabilitation staff.

Rehabilitation sessions (including in-person and virtual) are provided by respiratory therapists, clinical exercise physiologists, physical therapists, nurses, or others who have experience and training in either pulmonary or cardiac rehabilitation.

#### Appendix A: Control Intervention

Participants in the control arm receive two general education sessions and weekly phone call follow-ups during the Intervention Period. The first general education session is at the start of the Intervention Period, and the second is 2–3 weeks later. Weekly phone calls from site study staff will include education on topics of interest, including but not limited to fundamentals of exercise; self-management strategies; management of stress, anxiety, and depression; and nutrition and weight management. Participants are also asked to report the approximate time they spent exercising since the previous check-in.

#### Appendix B: Pacing Intervention

Pacing is a personalized strategy to prevent or reduce PEM, characterized by symptom exacerbation and prolonged recovery following activity. Pacing involves recognizing PEM episodes, identifying potential triggers (physical, orthostatic, cognitive, or emotional stressors), and determining individual activity limits (“energy envelope”) to balance activity and rest, thereby reducing symptom flares.^19^

Clinical sites are selected based on existing physical therapy, occupational therapy, rehabilitation therapy, or equivalent staff and services, and a pacing coach is selected who has:

- experience working with individuals with disability (e.g., Long COVID);
- familiarity with RECOVER-ENERGIZE training materials, including the pacing manual, participant workbook, supplementary videos, and completion of the training workshop.

Coaches receive ongoing group consultations for protocol adherence, professional support, and skill development. The pacing coach’s role is to lead the session, provide the participant with relevant education, build rapport, facilitate collaborative spirit, support the participant in applying the program strategies to manage PEM, and strategically use problem-solving skills to address barriers with the participant.

The intervention consists of 12 individual weekly video-conference sessions, each ∼30 minutes (25 minutes content, 5 minutes wrap-up). The goals of the structured pacing intervention are to 1) make a long-lasting change in the way that the participants think about PEM, including a discussion of common misconceptions of PEM as well as normalizing their experience of Long COVID; and 2) teach skills that participants can use to manage PEM, including pacing, activity logging, and energy conservation. Session 1 establishes expectations, and sessions 2–12 review pacing experiences, address questions, and reinforce skills. Topics include an introduction to structured pacing, activity analysis, rest and energy limits, introduction to pacing and activity logs, and creating a pacing plan. This structured, collaborative approach aims to improve recognition and response to PEM, maintain stable daily activity levels, and enhance predictability in quality of life.

#### Appendix B: Control Intervention

Usual care (i.e., care that the participant receives by their clinical team) is used for patients randomized to the control group. Participants in the control group receive basic education material about PEM and a weekly call for support and communication, but do not receive detailed guidance about pacing to prevent PEM. All study participants will be informed about the importance of adherence to study protocols and how participation, regardless of assignment to intervention or control, helps advance scientific knowledge.

### Study Activities

Both Appendix A (cardiopulmonary rehabilitation) and Appendix B (pacing) follow a structured schedule of assessments across a 6-month study period, consisting of screening, baseline, a 3-month intervention phase, and a 3-month follow-up phase. Procedures have been harmonized across the appendices where possible, while allowing for intervention-specific measures as depicted in **Figures 1 and 2**.

**Figure 1.**
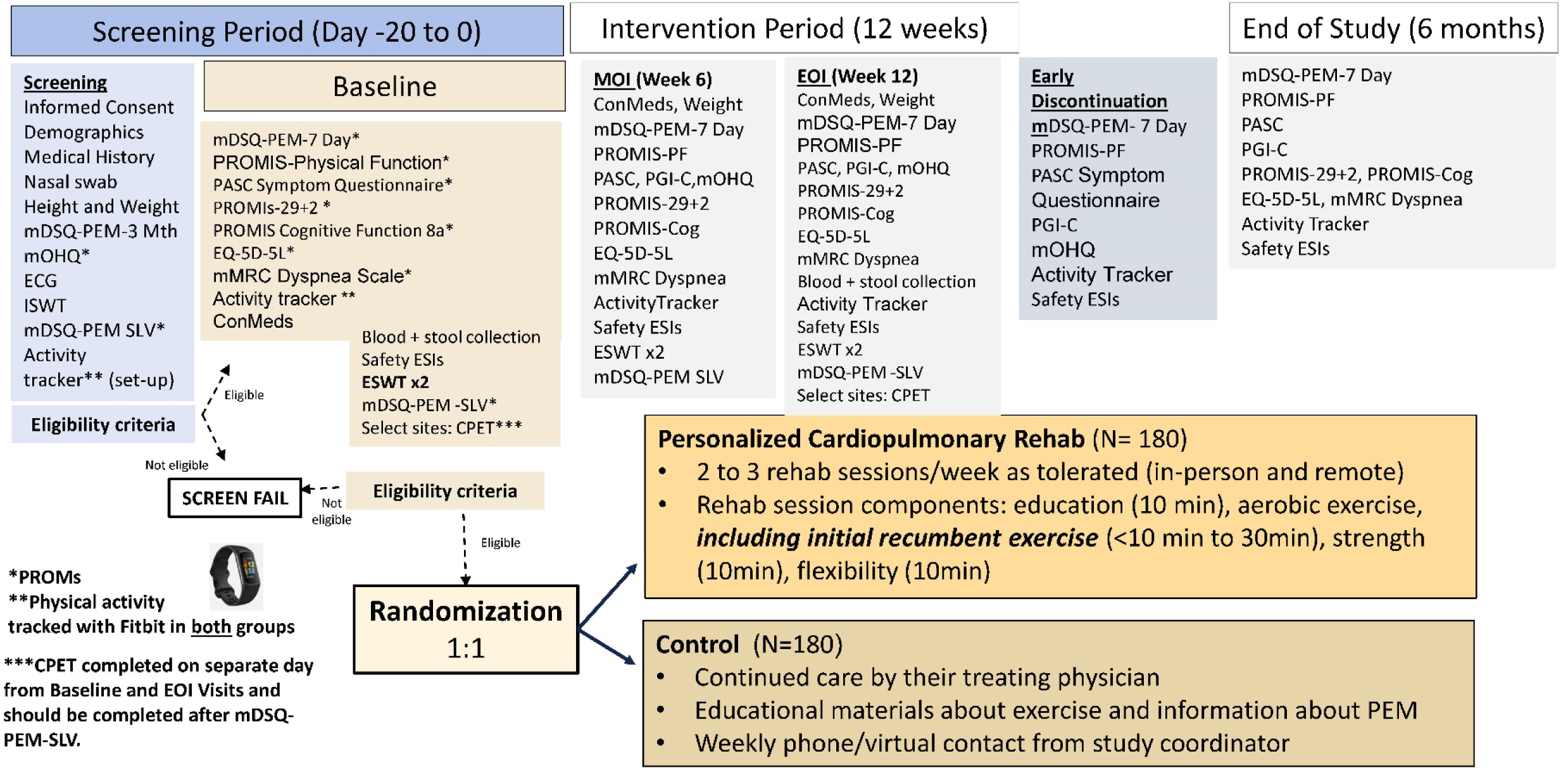
Appendix A (Cardiopulmonary Rehabilitation) Schema. Abbreviations: Cog: Cognitive Function Short Form 8a; CPET: cardiopulmonary exercise testing; ECG: electrocardiogram; EOI: end of intervention; EQ-5D-5L: EuroQol 5-Dimension 5-Level questionnaire; ESIs: events of special interest; ESWT: Endurance Shuttle Walk Test; ISWT: Incremental Shuttle Walk Test; mDSQ-PEM: modified DePaul Symptom Questionnaire–Post-Exertional Malaise; mMRC: modified Medical Research Council; mOHQ: Modified Orthostatic Hypotension Questionnaire; MOI: middle of intervention; Mth: month; PASC: post-acute sequelae of SARS-CoV-2 infection; PEM: post-exertional malaise; PF: Physical Function Short Form 8b; PGI-C: Patient Global Impression of Change; PROMIS: Patient-Reported Outcomes Measurement Information System; PROMs: patient-reported outcome measures; SLV: since last visit.

**Figure 2.**
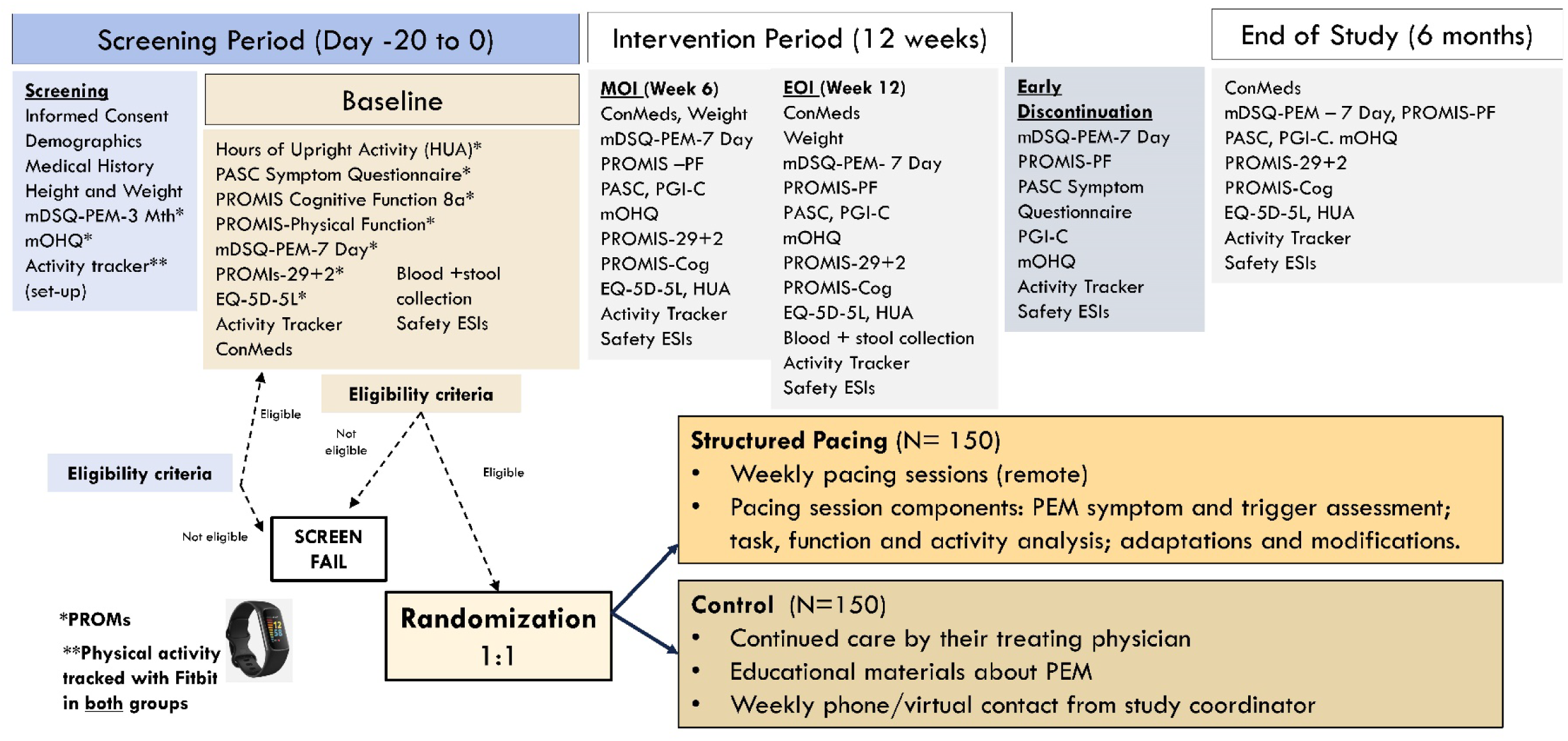
Appendix B (Pacing Intervention) Schema. Abbreviations: Cog: Cognitive Function Short Form 8a; EOI: end of intervention; EQ-5D-5L: EuroQol 5-Dimension 5-Level questionnaire; ESIs: events of special interest; HUA: hours of upright activity; mDSQ-PEM: modified DePaul Symptom Questionnaire–Post-Exertional Malaise; mOHQ: Modified Orthostatic Hypotension Questionnaire; MOI: middle of intervention; Mth: month; PASC: post-acute sequelae of SARS-CoV-2 infection; PEM: post-exertional malaise; PF: Physical Function Short Form 8b; PGI-C: Patient Global Impression of Change; PROMIS: Patient-Reported Outcomes Measurement Information System; PROMs: patient-reported outcome measures.

### Primary Outcome Measures

#### Appendix A

The ESWT measures exercise capacity via timed walking on a 10-meter course.^24^ Results are total walking time following a 2-minute warm-up. Baseline walking speed is determined by the ISWT, which increases pace each minute until participants can no longer continue. The ESWT is performed at 85% of maximum ISWT speed, twice, at least 30 minutes apart; the longer time is used for analyses.

#### Appendix B

The mDSQ-PEM assesses PEM and is adapted to include different look-back periods (e.g., past 3 months, past 7 days, or since last visit) to track symptom change during the study.^23^ Outcomes include frequency, duration, and severity scores to evaluate intervention effects rather than using the measure as a rule-out, rule-in tool.

### Additional Outcome Measures

Cardiopulmonary exercise testing, specifically symptom-limited maximal treadmill or cycle test (2–3 minutes warm-up, 8–12 minutes increasing work rate), is used at select sites for Appendix A. The primary measure is peak oxygen uptake (VO₂ peak).

Also, activity tracker devices (e.g., Fitbit®) are to be worn 24 hours per day for ≥7 days before baseline and throughout the study for both appendices. Metrics include average daily step count. For Appendix B participants, activity trackers are used as a tool for the pacing interventions by monitoring heart rate and step thresholds to prevent PEM. Successful pacing is expected to stabilize or reduce step counts, minimizing symptom flares rather than increasing activity.

### Statistical Analyses

#### Sample Size and Power

The recruitment goals of 360 patients for Appendix A and 300 patients for Appendix B, randomized 1:1 to study interventions versus controls, are based on the following considerations: (1) expected loss to follow-up to be up to 20% and (2) statistical power to exceed 85% under conservative assumptions for the expected treatment effects and variability for the primary endpoints. For Appendix A, the primary endpoint is change in ESWT, with adjustment for baseline ESWT. Expected effect sizes and variability were determined using estimates from Daynes et al. (2021),^26^ which included a baseline mean ESWT estimate of 300 seconds, a range of absolute improvements (treatment effects) of 150–300 seconds, a range of variability for the SDs of 400–500 seconds, and a range of correlations between baseline and follow-up ESWT of 0.3–0.7. For Appendix B, the primary endpoints are the mean change in the PEM symptoms (mean frequency scores, mean severity scores, and duration) using the mDSQ-PEM questionnaire. Expected treatment effects and variability were conservatively estimated using published estimates from Jason et al.^27^ We consider treatment effect differences of 0.4 points and a conservatively inflated SD of 1.15 points. Jason et al. reported SDs between 1.08–1.11 for individuals comparing their ME/CFS treatment to controls. For Appendix B, power was estimated using an independent two-sample t-test for the frequency and severity outcomes and an ordered logit model for the duration outcome. We assumed all tests were two-sided with a .05 significance level.

### Primary Analyses

#### Appendix A

For this study, data from all enrolled participants who receive the control intervention or treatment intervention will be included in the analyses. Between-group comparisons of the primary study endpoint, the change from baseline to end of intervention in ESWT, will be analyzed using linear regression, adjusting for the baseline ESWT as a concomitant variable and possibly prespecified baseline characteristics (details will be provided in the statistical analysis plan). Model standard errors will be robustly estimated using Huber–White (sandwich) standard errors. The mean difference between the treatment groups for the primary endpoint will be compared for the modified intention-to-treat population.

#### Appendix B

Data from all randomized participants who receive the control intervention or pacing intervention will be included in the analyses. We will assess between-group comparisons of the primary study endpoints, the change from baseline to end of intervention in the mDSQ-PEM mean frequency score, and mean severity score using linear regression, adjusting for their respective pretreatment values as adjustment variables. We will analyze duration using an ordered logit regression model.

For some secondary and exploratory endpoints (e.g., PROMIS Cognitive Function Short Form 8a, PROMIS-29+2, PROMIS Physical Function Short Form 8b, and activity tracker step count), we will use linear regression models similar in form to the regression models used for the primary endpoints for independent data. For repeated-measures data, we will use generalized estimating equation regression models that account for repeated-measures correlation, to evaluate treatment differences. For other endpoints (PASC Symptom Questionnaire, EuroQol 5-Dimension 5-Level questionnaire [EQ-5D-5L], and mOHQ), we will analyze via descriptive statistics. Please see **Tables 1 and 2** for a complete list of outcomes for Appendix A and Appendix B. For a detailed presentation of the statistical approaches used for the primary study endpoints and secondary analyses, the reader is referred to the platform protocol available at https://clinicaltrials.gov/study/NCT06404047. All hypothesis tests and confidence interval estimates will be two-sided. A Type I error rate of 0.05 will be used to investigate the hypothesis of the primary objective.

**Table 1.** Appendix A (Cardiopulmonary Rehabilitation) Objectives and Outcome Measures.

| Objective | Outcome Measure |
| --- | --- |
| Primary |  |
| Evaluate the effect of study intervention on exercise tolerance versus control<sup>a</sup> | Endurance Shuttle Walk Test |
| Secondary |  |
| Efficacy – Evaluate the effect of study intervention on physical function versus control<sup>b</sup> | PROMIS Physical Function Short Form 8b, step count |
| Safety – Evaluate the effect of study intervention on PEM symptoms (frequency, severity, and duration) versus control<sup>b</sup> | Modified DePaul Symptom Questionnaire–Post-Exertional Malaise |
| Exploratory |  |
| Evaluate the effect of study intervention on shortness of breath, cognitive function, quality of life, and Long COVID symptoms versus control<sup>b</sup> | PASC Symptom Questionnaire, Modified Medical Research Council Dyspnea Scale, PROMIS Cognitive Function Short Form 8a, PROMIS-29+2, EQ-5D-5L |
| Evaluate the effect of study intervention on orthostatic hypotension versus control<sup>c</sup> | Modified Orthostatic Hypotension Questionnaire |
| Evaluate the effect of study intervention on relevant biomarkers versus control<sup>a</sup> | Viral biomarkers, immune biomarkers |
| Evaluate the effect of study intervention on healthcare utilization versus control<sup>d</sup> | Healthcare utilization – medical visits and hospitalizations |
| Describe the effects of study intervention on exercise capacity (at select sites)<sup>e</sup> | Cardiopulmonary exercise testing |
| Compare the effect of study intervention on physical function versus control<sup>b</sup> | Actigraphy – heart rate |

<sup>a</sup>Endpoint: change from baseline to end of intervention in the treatment group compared to control group.
<sup>b</sup>Endpoint: change from baseline to the middle of intervention, end of intervention, and end of study in the treatment group compared to control group.
<sup>c</sup>Endpoint: change from screening to the middle of intervention, end of intervention, and end of study in the treatment group compared to control group.
<sup>d</sup>Endpoint: cumulative medical visits or hospitalizations from baseline to end of intervention or end of study.
<sup>e</sup>Endpoint: change from baseline to end of intervention in physiological responses to cardiopulmonary exercise testing.
EQ-5D-5L: EuroQol 5-Dimension 5-Level questionnaire; PASC: post-acute sequelae of SARS-CoV-2 infection; PEM: post-exertional malaise; PROMIS: Patient-Reported Outcomes Measurement Information System.

**Table 2.** Appendix B (Pacing Intervention) Objectives and Outcome Measures.

| Objective | Outcome Measure |
| --- | --- |
| Primary |  |
| Evaluate the effect of study intervention on PEM symptoms (frequency, severity, and duration) versus control<sup>a</sup> | Modified DePaul Symptom Questionnaire–Post-Exertional Malaise |
| Secondary |  |
| Evaluate the effect of study intervention on Long COVID symptoms versus control<sup>b</sup> | PROMIS Cognitive Function Short Form 8a |
| Evaluate the effect of study intervention on QoL versus control<sup>b</sup> | PROMIS-29+2, EQ-5D-5L |
| Evaluate the effect of study intervention on physical activity versus control<sup>b</sup> | Activity trackers (Fitbit®), PROMIS Physical Function Short Form 8b |
| Evaluate the effect of study intervention on orthostatic intolerance versus control<sup>b</sup> | Modified Orthostatic Hypotension Questionnaire |
| Exploratory |  |
| Evaluate the effect of study intervention on physical activity versus control<sup>b</sup> | Self-reported hours of upright activity on best day and on worst day in last week |
| Evaluate the effect of study intervention on relevant biomarkers versus control<sup>a</sup> | Viral biomarkers, immune biomarkers |
| Evaluate the effect of study intervention on healthcare utilization versus control<sup>a</sup> | Healthcare utilization – medical visits and hospitalizations |
| Evaluate the effect of study intervention on Long COVID symptoms versus control<sup>b</sup> | PASC Symptom Questionnaire |

<sup>a</sup>Endpoint: change from baseline to end of intervention in the treatment group compared to control group.
<sup>b</sup>Endpoint: change from baseline to the middle of intervention, end of intervention, and end of study in the treatment group compared to control group.
EQ-5D-5L: EuroQol 5-Dimension 5-Level questionnaire; PASC: post-acute sequelae of SARS-CoV-2 infection; PROMIS: Patient-Reported Outcomes Measurement Information System.

### Planned Interim Analyses

Interim examination of serious adverse events will be performed at regular intervals during the course of the trial. An independent, NIH-appointed data and safety monitoring board will monitor participant safety and review participant enrollment and performance of the trial. The primary objective of these interim analyses will be to ensure the safety of the participants enrolled in the trial and evaluate the accumulating endpoint data by treatment group. In addition, interim monitoring will involve a review of participant recruitment, compliance with the study protocol, status of data collection, and other factors that reflect the overall progress and integrity of the study.

The protocol does not include plans for early stopping based on efficacy. The wide variability of Long COVID symptoms and outcomes underscores the importance of estimating treatment effects across a comprehensive range of participant-relevant measures. Stopping the study before full enrollment would diminish analytic precision, hinder assessment of key secondary and safety outcomes, and restrict the evidence base needed to inform subsequent clinical trials in this population.

### Patient and Public Involvement

When the NIH formed the RECOVER Initiative in February 2021 to support research focused on understanding and treating Long COVID, it created a robust structure that included patients, caregivers, and community representatives to ensure that RECOVER-CT would meet the needs of people affected by this chronic condition. Representatives who had participated in prior RECOVER observational activities or reviewed the intervention and study proposals submitted by investigators to the NIH for the clinical trials were invited to be part of the protocol development process for ENERGIZE. The design of the trial was iteratively developed, incorporating critical input from the patient representatives. These representatives were instrumental in the development of the concept for Appendix B to address the concerns related to possible inclusion of patients with PEM in a cardiopulmonary rehabilitation intervention. The choice of the structured pacing intervention, the inclusion and exclusion criteria for both Appendices, and the outcome measures used, particularly to assess for PEM, were developed in close coordination with our patient representatives.

## DISCUSSION

Exercise intolerance is one of the most prevalent and disabling symptoms in Long COVID, and is often accompanied by PEM. No large randomized controlled trials have yet evaluated interventions targeting both symptoms. The RECOVER-ENERGIZE platform trial is a prospective, multicenter, phase 2 randomized study comparing two approaches: cardiopulmonary rehabilitation for participants without significant PEM, and structured pacing for those with moderate to severe PEM.

Traditional cardiopulmonary rehabilitation benefits many with cardiopulmonary disease, but Long COVID requires adaptation due to orthostatic intolerance and PEM risk. Exercise may exacerbate PEM; therefore, systematic PEM screening and monitoring with the mDSQ-PEM is an integral and novel feature of this trial. Administered at baseline, the mDSQ-PEM directs participants to the appropriate appendix (A or B) and is repeated throughout the trial to detect new or worsening PEM, enabling timely modification of interventions. In the pacing trial, the mDSQ-PEM serves as a primary outcome measure, assessing severity, frequency, and duration of PEM. Critically, the rehabilitation intervention in Appendix A is not graded exercise therapy; it is an individualized, symptom-limited program with built-in safeguards. Participants who develop significant PEM during the intervention are discontinued from Appendix A and may be offered enrollment in Appendix B. Similarly, individuals who screen positive for significant PEM during the Appendix A screening process are directed to Appendix B. These design features ensure participant safety while maintaining the opportunity for all eligible participants to receive an appropriate intervention.

The structured pacing intervention, modeled on ME/CFS management, is an innovative feature given its use of individualized, one-on-one coaching for Long COVID. Delivered in-person or remotely, with flexible short sessions and repeated content for cognitive or fatigue-related impairments, pacing aims to stabilize symptoms and prevent the “push–crash cycle.” Activity levels are expected to stabilize rather than increase. Upright activity hours are also assessed in this trial, reflecting patient-advocate priorities identified during the design phase of the trial.

Scalability was a deliberate design consideration. Pacing coaches are required to have rehabilitation clinical experience but may come from diverse disciplines, including medical assistants, nurses, clinical exercise physiologists, occupational therapists, and physical therapists. We intentionally allowed for a broad range of background experience when choosing pacing coaches to allow for scalability of the intervention across a variety of different types of health systems and settings. Centralized training led by clinicians experienced in delivering behavioral interventions for fatigue management, coupled with standardized materials, is also intended to facilitate broad dissemination should the intervention prove effective.

Cardiopulmonary rehabilitation programs are present in many medical centers. If rehabilitation is found to be an effective treatment for exercise intolerance in Long COVID, existing rehabilitation programs may be scaled up to treat individuals with Long COVID to appropriately evaluate and manage PEM. In addition, the results of this trial may warrant creation of new programs in the community to address these disabling symptoms.

If successful, RECOVER-ENERGIZE will provide evidence for two symptom-targeted interventions based on PEM status: rehabilitation to enhance exercise capacity and quality of life in those without PEM, and pacing to reduce PEM severity and improve functional stability in those with PEM. The trial’s focus on safety, tailored interventions, and scalability supports rapid integration into routine care, with potential to substantially reduce symptom burden and improve quality of life for individuals with Long COVID.

## Ethics and Dissemination

The RECOVER-ENERGIZE trial protocol has been approved by an institutional review board (Advarra), and written informed consent will be obtained from all participants prior to enrollment. The trial is registered on ClinicalTrials.gov (NCT06404047). Formally assessing PEM and developing a structured activity pacing intervention delivered by local pacing coaches are novel features of this trial. Results will be disseminated through peer-reviewed publications, presentations at scientific conferences, and communication with participants, patient advocacy organizations, and the broader Long COVID community. De-identified participant data will be made available through the NIH RECOVER data repository in accordance with NIH data-sharing policies. If successful, this protocol will provide accessible tools that clinicians can use to address exercise intolerance and PEM in patients with Long COVID.

## Data Availability

De-identified participant data will be made available through the NIH RECOVER data repository in accordance with NIH data-sharing policies.

## Acknowledgments

We acknowledge Amy Patterson, MD, and Antonello Punturieri, MD, from the National Heart, Lung, and Blood Institute; Michael Proschan, PhD, from the National Institute of Allergy and Infectious Diseases; and Frank C. Sciurba, MD, from the University of Pittsburgh School of Medicine for their assistance with oversight and review of the protocol, as well as Peter Hoffmann of the Duke Clinical Research Institute (DCRI) who provided assistance with editing and submission of the manuscript, as part of his employment with DCRI.

## Author contributions

J.F. drafted the initial manuscript and takes responsibility for the integrity of the data collection, accuracy, and its analysis. All authors contributed to the study design and drafting of the manuscript. All authors have read and approved the final version of the manuscript. The corresponding author attests that all listed authors meet authorship criteria and that no others meeting the criteria have been omitted.

