## Supplemental Material for "Rationale and Design of RECOVER-ENERGIZE: A Platform Clinical Trial of Interventions for Exercise Intolerance With and Without Post-exertional Malaise in Long COVID"

### DePaul Symptom Questionnaire – Post Exertional Malaise short form (DSQ-PEM)

Subject ID: ______________________ Site: ________ Date: __________ Visit: ______________________

For each symptom below, please circle one number for frequency and one number for severity: Please complete the chart from left to right.

| Symptoms | Frequency: Throughout the **past 3 months**, **how often** have you had this symptom? For each symptom listed below, circle a number from: **0 = none of the time** **1 = a little of the time** **2 = about half the time** **3 = most of the time** **4 = all of the time** | | | | | Severity: Throughout the **past 3 months**, **how much** has this symptom bothered you? For each symptom listed below, circle a number from: **0 = symptom not present** **1 = mild** **2 = moderate** **3 = severe** **4= very severe** | | | | |
|---|---|---|---|---|---|---|---|---|---|---|
| 1. Dead, heavy feeling after starting to exercise | 0 | 1 | 2 | 3 | 4 | 0 | 1 | 2 | 3 | 4 |
| 2. Next day soreness or fatigue after non-strenuous, everyday activities | 0 | 1 | 2 | 3 | 4 | 0 | 1 | 2 | 3 | 4 |
| 3. Mentally tired after the slightest effort | 0 | 1 | 2 | 3 | 4 | 0 | 1 | 2 | 3 | 4 |
| 4. Minimum exercise makes you physically tired | 0 | 1 | 2 | 3 | 4 | 0 | 1 | 2 | 3 | 4 |
| 5. Physically drained or sick after mild activity | 0 | 1 | 2 | 3 | 4 | 0 | 1 | 2 | 3 | 4 |

For each question below, choose the answer which best describes your PEM symptoms.

| | | | | | | |
|---|---|---|---|---|---|---|
| 6. If you were to become exhausted after actively participating in extracurricular activities, sports, or outings with friends, would you recover within an hour or two after the activity ended? | Yes | | | No | | |
| 7. Do you experience a worsening of your **fatigue/energy related illness** after engaging in minimal **physical** effort? | Yes | | | No | | |
| 8. Do you experience a worsening of your **fatigue/energy related illness** after engaging in minimal **mental** effort? | Yes | | | No | | |
| 9. If you feel worse after activities, how long does this last? | ≤1 h | 2-3 h | 4-10 h | 11-13 h | 14-23 h | ≥24 h |
| 10. If you do not exercise, is it because exercise makes your symptoms worse? | Yes | | | No | | |

Used with permission of Leonard A. Jason, PhD.
